# Plasma MIA, CRP, and albumin predict cognitive decline in Parkinson’s Disease

**DOI:** 10.1101/2022.03.16.22272456

**Authors:** Junchao Shen, Noor Amari, Rebecca Zack, R. Tyler Skrinak, Travis L. Unger, Marijan Posavi, Thomas F. Tropea, Sharon X. Xie, Vivianna M. Van Deerlin, Richard B. Dewey, Daniel Weintraub, John Q. Trojanowski, Alice S. Chen-Plotkin

## Abstract

**Objective:** Using a multi-cohort, Discovery-Replication-Validation design, we sought new plasma biomarkers that predict which PD individuals will experience cognitive decline.

**Methods:** In 108 Discovery Cohort PD individuals and 83 Replication Cohort PD individuals, we measured 940 plasma proteins on an aptamer-based platform. Using proteins associating with subsequent cognitive decline in both cohorts, we trained a logistic regression model to predict which PD patients showed fast (>=1 point drop/year on Montreal Cognitive Assessment (MoCA)) vs. slow (<1 point drop/year on MoCA) cognitive decline in the Discovery Cohort, testing it in the Replication Cohort. We developed alternate assays for the top three proteins and confirmed their ability to predict cognitive decline – defined by change in MoCA or development of incident Mild Cognitive Impairment (MCI) or dementia – in a Validation Cohort of 118 PD individuals. We investigated the top plasma biomarker for causal influence by Mendelian randomization.

**Results:** A model with only three proteins (Melanoma Inhibitory Activity Protein (MIA), C-Reactive Protein (CRP), albumin) separated Fast vs. Slow cognitive decline subgroups with an AUC of 0.80 in the Validation Cohort. Validation Cohort PD individuals in the top quartile of risk for cognitive decline based on this model were 4.4 times more likely to develop incident MCI or dementia than those in the lowest quartile. Genotypes at *MIA* SNP rs2233154 associated with MIA levels and cognitive decline, providing evidence for MIA’s causal influence.

**Conclusions:** An easily-obtained plasma-based predictor identifies PD individuals at risk for cognitive decline. MIA may participate causally in development of cognitive decline.

## INTRODUCTION

Parkinson’s disease (PD) is the second most common neurodegenerative disease affecting more than 5 million people worldwide. While the progressive loss of dopaminergic neurons in the substantia nigra pars compacta (SNc) results in PD motor symptoms of bradykinesia, tremor, and rigidity,^1^ cognitive impairment and dementia also develop in a large proportion of individuals with PD,^2^ exacting a high financial and emotional cost for patients, their families, and the healthcare system.^3,4,5^ Among PD patients, there is marked heterogeneity in cognitive trajectory, making prognostication difficult and creating barriers for clinical trials aimed at modifying this important aspect of disease.^6^ As a consequence, biomarkers predictive of cognitive decline in PD are urgently needed, particularly if they might also shed light on the mechanisms underlying development of cognitive decline in some PD patients but not others.

To date, such biomarkers of PD cognitive decline are sparse.^7^ In the cerebrospinal fluid (CSF), higher phosphorylated tau and lower amyloid β_42_ are associated with a higher risk of dementia in PD,^8,9^ and higher neurofilament light chain (NfL) also predicts cognitive decline.^10^ In the plasma, both higher NfL and lower epidermal growth factor levels predict cognitive decline in PD.^11,12,13,14,15^ Extracellular-vesicle-associated tau and amyloid β_42_ have also been reported to correlate with cognition in PD.^16^ Additionally, genetic variants have been linked to cognitive trajectory in PD,^17,18,19^ with the most well-replicated effects on cognition seen for the *APOE* E4 allele^20,21,22,23^ and PD-associated *GBA* variants.^17,24,25,26^

Studies to date, however, have limitations that hamper translation of biomarkers of PD cognitive decline into clinical contexts. First, while some biomarkers have been widely replicated, emerging biomarkers are often described in relatively small cohorts without replication and validation.^27^ Second, statistical associations may need to be converted into risk scores or other tools that can be meaningfully used for risk stratification at the individual level. Third, and perhaps most difficult to address, biomarkers emerging from large-scale screens (vs. biomarkers tested for reasons related to, for example, their known role in Alzheimer’s disease pathogenesis or *GBA*-related pathways) often lack biological context, leading to difficulty discerning whether a given biomarker candidate is simply correlated with a phenotype or causally involved in its development.^28^

Here, we aimed to address these gaps by utilizing a multi-cohort, multi-stage design starting with a screen of 940 plasma proteins in 191 longitudinally-followed PD patients. From these initial data, we validated a three-protein blood-based biomarker panel in an additional 118 longitudinally-followed PD patients, demonstrating that this protein panel enriches for PD individuals who will experience rapid cognitive decline regardless of cohort studied, cognitive measure used, or method of biomarker measurement. Finally, we perform Mendelian-randomization-based analyses to probe one of these newly-discovered biomarkers for evidence of causality.

## METHODS

### Overview of study design

108 PD patients were enrolled at the University of Texas Southwestern (UTSW Discovery Cohort) as part of the NIH-NINDS Parkinson’s Disease Biomarker Program (PDBP). 83 PD patients were enrolled at the University of Pennsylvania (UPenn Replication Cohort). The Discovery and Replication Cohorts were screened for 1,129 and 1,305 proteins, respectively, using an aptamer-based platform assay called SOMAScan.^29^ A total of 940 proteins that passed quality control (QC) metrics, as previously described,^30^ in both cohorts, were retained for further analysis. We then (1) assigned individuals to fast vs. slow cognitive decline subgroups based on the rate of decline in MoCA score, and (2) identified which proteins differentiated fast vs. slow cognitive decline subgroups in the Discovery and Replication Cohorts, to (3) develop a multi-protein model for predicting cognitive course in PD, before (4) validating top biomarker candidates with alternative assays (ELISA and BCP), and (5) testing the final predictive model in a Validation Cohort (118 patients from University of Pennsylvania, **Fig. 1)**.

**Fig 1.**
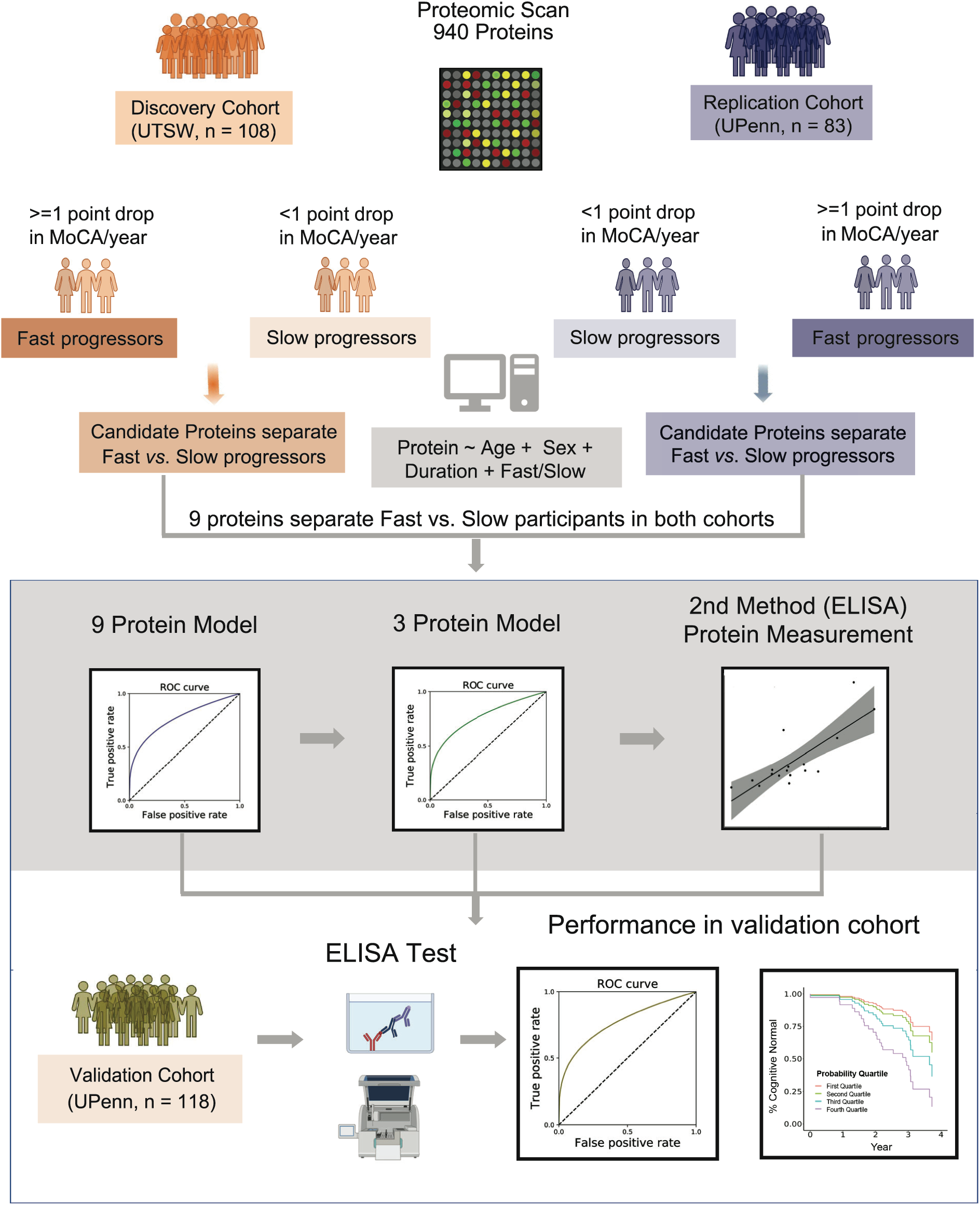
Study overview. An aptamer-based platform was used to quantify the plasma levels of 940 proteins in the Discovery Cohort (left panel) and Replication Cohort (right panel). In each cohort, PD patients were assigned into a fast or a slow cognitive decline group based on change in MoCA score over time. A linear regression model was used to identify proteins differentiating fast versus slow progressors in both cohorts, generating the top 9 proteins. Next, these identified proteins were used to train two logistic regression-based models that predict whether individual PD patients subsequently have fast vs. slow cognitive decline. Finally, in an additional validation cohort of 118 PD patients, we measured top biomarker proteins using alternative assays, testing for their performance in separating fast vs. slow cognitive decline subgroups.

### Cohorts and sample collection

A total of 309 PD participants who were non-demented at the baseline visit, with blood samples collected between 2013 and 2019 through the University of Pennsylvania (UPenn) and NIH-NINDS Parkinson’s Disease Biomarker Program (PDBP, with UTSW as the collection site) were included in the analysis.^31,32^ All individuals met diagnostic criteria of the United Kingdom Parkinson’s Disease Brain Bank for PD. Demographics are summarized in **Table 1**, and details are in the Supplementary Methods.

**Table 1.**
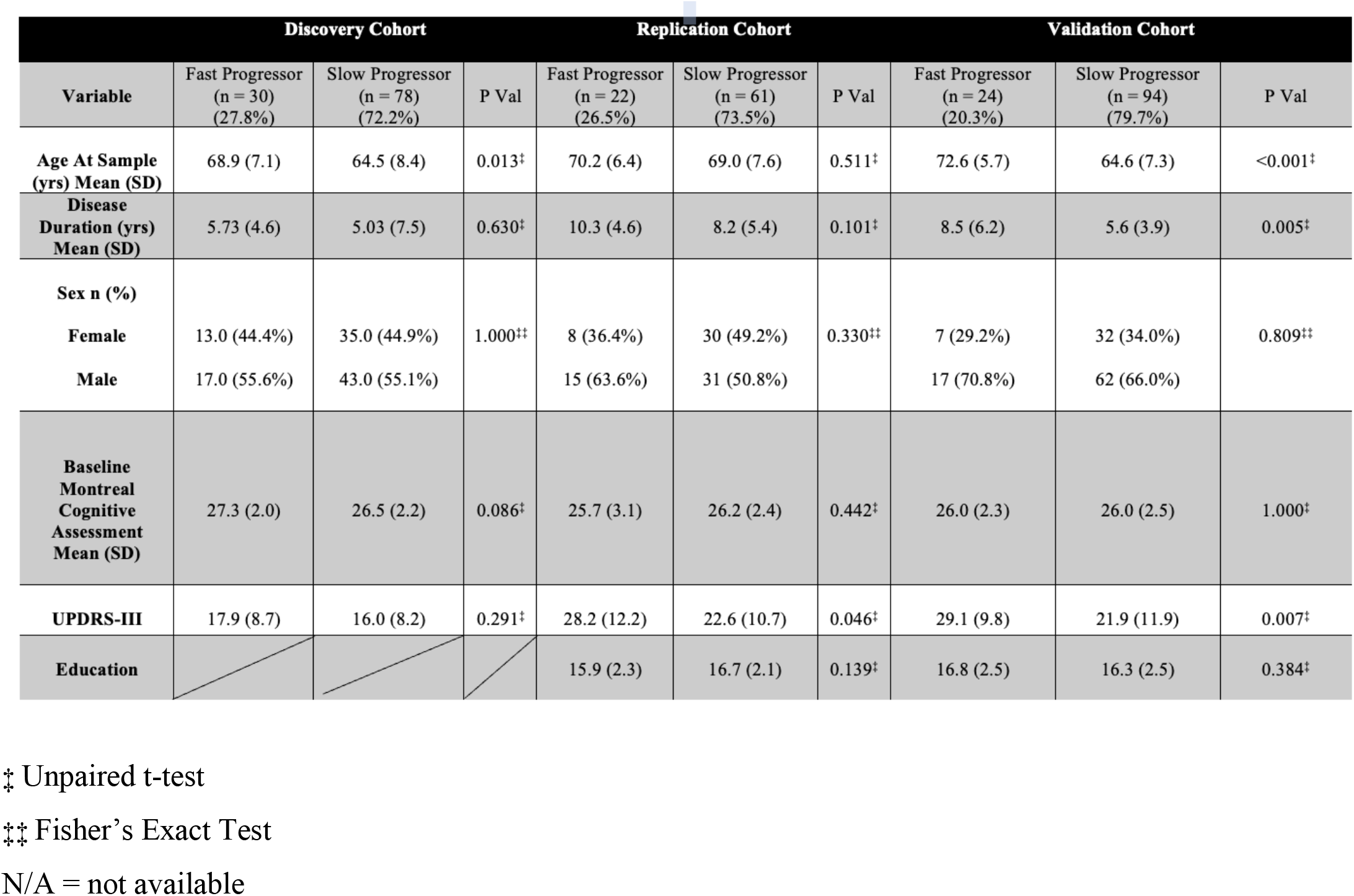
Demographic characteristics of the Discovery (UTSW), Replication (UPenn), and Validation (UPenn) cohorts. Within each cohort, individuals defined as having a faster rate of cognitive decline vs. a slower rate of cognitive decline based on MoCAscore change over time are compared as indicated with the symbol, based on distribution of the data. Years of education were not available for the Discovery Cohort.

**TABLES AND FIGURE LEGENDS**
| Variable | Discovery Cohort |  |  | Replication Cohort |  |  | Validation Cohort |  |  |
| --- | --- | --- | --- | --- | --- | --- | --- | --- | --- |
|  | Fast Progressor<br>(n = 30)<br>(27.8%) | Slow Progressor<br>(n = 78)<br>(72.2%) | P Val | Fast Progressor<br>(n = 22)<br>(26.5%) | Slow Progressor<br>(n = 61)<br>(73.5%) | P Val | Fast Progressor<br>(n = 24)<br>(20.3%) | Slow Progressor<br>(n = 94)<br>(79.7%) | P Val |
| Age At Sample<br>(yrs) Mean (SD) | 68.9 (7.1) | 64.5 (8.4) | 0.013<sup>‡</sup> | 70.2 (6.4) | 69.0 (7.6) | 0.511<sup>‡</sup> | 72.6 (5.7) | 64.6 (7.3) | <0.001<sup>‡</sup> |
| Disease<br>Duration (yrs)<br>Mean (SD) | 5.73 (4.6) | 5.03 (7.5) | 0.630<sup>‡</sup> | 10.3 (4.6) | 8.2 (5.4) | 0.101<sup>‡</sup> | 8.5 (6.2) | 5.6 (3.9) | 0.005<sup>‡</sup> |
| Sex n (%) |  |  |  |  |  |  |  |  |  |
| Female | 13.0 (44.4%) | 35.0 (44.9%) | 1.000<sup>‡‡</sup> | 8 (36.4%) | 30 (49.2%) | 0.330<sup>‡‡</sup> | 7 (29.2%) | 32 (34.0%) | 0.809<sup>‡‡</sup> |
| Male | 17.0 (55.6%) | 43.0 (55.1%) |  | 15 (63.6%) | 31 (50.8%) |  | 17 (70.8%) | 62 (66.0%) |  |
| Baseline<br>Montreal<br>Cognitive<br>Assessment<br>Mean (SD) | 27.3 (2.0) | 26.5 (2.2) | 0.086<sup>‡</sup> | 25.7 (3.1) | 26.2 (2.4) | 0.442<sup>‡</sup> | 26.0 (2.3) | 26.0 (2.5) | 1.000<sup>‡</sup> |
| UPDRS-III | 17.9 (8.7) | 16.0 (8.2) | 0.291<sup>‡</sup> | 28.2 (12.2) | 22.6 (10.7) | 0.046<sup>‡</sup> | 29.1 (9.8) | 21.9 (11.9) | 0.007<sup>‡</sup> |
| Education |  |  |  | 15.9 (2.3) | 16.7 (2.1) | 0.139<sup>‡</sup> | 16.8 (2.5) | 16.3 (2.5) | 0.384<sup>‡</sup> |

<sup>‡</sup> Unpaired t-test
<sup>‡‡</sup> Fisher's Exact Test
N/A = not available

### Protein quantification

Plasma proteins were quantified by SOMAscan in the Discovery and Replication Cohorts, then validated by ELISA or BCP assay in the Validation Cohort.

#### SOMAScan

Plasma samples from both the Discovery and Replication Cohorts were assayed using the 1.1k and 1.3k Assay versions of the SOMAScan platform (Somalogic, Boulder, CO, USA) – based on aptamer capture of protein targets – as previously described.^30^ SOMAScan proteins that did not meet previously-described quality control metrics^30^ were eliminated, leaving 940 plasma protein candidates. Protein measures are in relative fluorescence units (RFUs), log_10_ transformed for downstream analyses.

#### Enzyme-linked immunosorbent assay (ELISA)

For C-reactive protein, or CRP, and melanoma inhibitory activity protein, or MIA, ELISAs were used in the Validation Cohort. For CRP, we used the Human CRP Quantikine ELISA Kit (R&D CDRP00), diluting plasma samples 1:200. For MIA, we used the MIA ELISA Kit (Roche 11976826001), diluting plasma samples 1:2.

#### Bromocresol Purple Assay (BCP)

For albumin, we employed the widely-used bromocresol purple assay, using the BCP Albumin Assay Kit (Sigma MAK125), diluting plasma samples 1:5.

### Comparison of protein measures across assay platforms

For 15 individuals, we obtained two different aliquots of plasma sampled and banked on the same day. For one aliquot, proteins were assayed by SOMAScan. For the other aliquot, proteins were assayed by MIA ELISA, CRP ELISA, and BCP albumin assay. We compared measures for these 15 duplicate samples across platforms, obtaining Pearson’s r and p-values for correlation.

### Categorization of PD participants into fast and slow cognitive decline groups

Participants in the Discovery, Replication, and Validation Cohorts were categorized into two subgroups (Fast vs. Slow Cognitive Decline) based on rates of change in MoCA score. The rate of MoCA decline was calculated using the MoCA score difference between the patient’s last follow-up visit and baseline visit divided by duration of time (years) between the two visits [i.e., (MoCA at last visit -MoCA at baseline visit)/years)]. Patients with a MoCA score decline rate of 1 or more points/year were categorized as a fast cognitive decline subgroup, while the remaining patients were assigned to a slow cognitive decline subgroup.

### Statistical Analyses

All analyses were performed in R. R-scripts are available in the Supplement, along with details of statistical analyses.

### Mendelian randomization-based analyses of MIA

We used Mendelian randomization (MR)^33^ to test the hypothesis that the top biomarker MIA is causally related to development of cognitive decline in PD. Querying the Genotype-Tissue Expression (GTEx) database,^34^ we identified 4 linked single nucleotide polymorphisms (SNPs) at the *MIA* locus demonstrating significant expression quantitative locus (eQTL) effects. We tested these for association with plasma MIA protein levels, finding that all were protein expression quantitative trait loci (pQTLs), with rs2233154 showing the strongest correlation.

Next, we tested for association between rs2233154 genotypes and cognitive decline in 180 PD patients (all individuals from Replication and Validation Cohorts combined for which genotyping data was available). We assessed the effect of rs2233154 genotype on cognitive change over five years of follow-up using linear mixed-effects models as well as Cox proportional hazards models adjusted for age, sex, and disease duration.

## RESULTS

### Subgroups of Parkinson’s Disease differing by rate of cognitive decline

In 108 longitudinally-followed PD patients from the UTSW-based Discovery Cohort (**Table 1**), 30 individuals (28%) declined by 1 or more points per year on the MoCA and were assigned to the Fast Cognitive Decline subgroup. In 83 longitudinally-followed PD patients from the UPenn-based Replication Cohort (**Table 1**), 22 individuals (27%) were assigned to the Fast Cognitive Decline subgroup. Remaining PD patients were assigned to the Slow Cognitive Decline subgroup.

To ensure that age, sex, or disease duration was not driving the rate of cognitive decline in the Fast vs. Slow Cognitive Decline subgroups, we compared these subgroups in both the Discovery and Replication Cohorts using linear mixed-effects models adjusted for these variables. As shown in **Figures 2A** and **2B**, Fast and Slow Cognitive Decline subgroups differed significantly in rate of change in MoCA even after adjustment for age, sex, and disease duration (p<0.001 for both cohorts).

**Fig 2.**
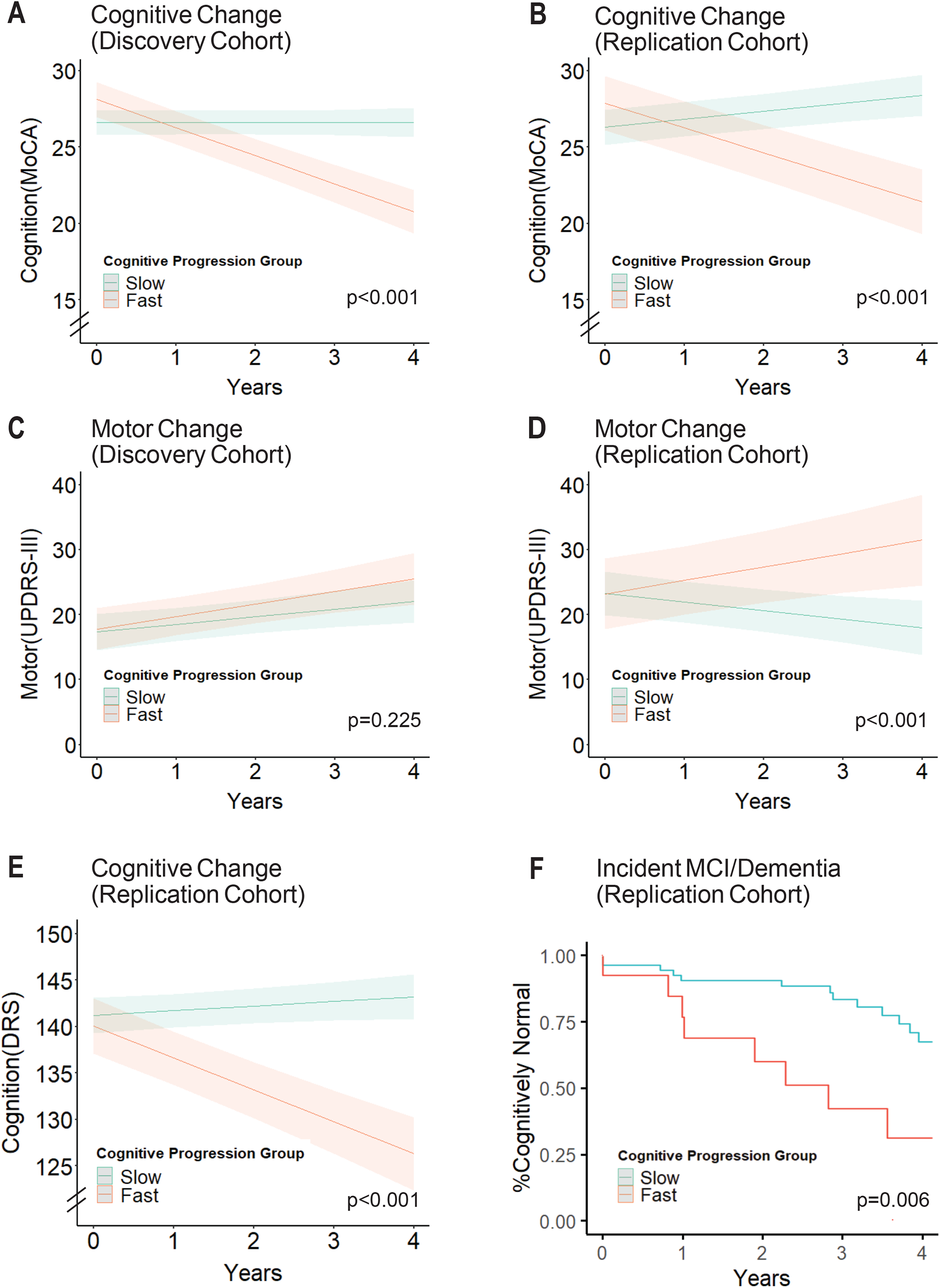
Characterization of cognitive decline subgroups. Longitudinal cognitive and motor performance in the fast versus slow cognitive decline subgroups was assessed using linear mixed-effect models adjusting for age, sex and disease duration. Subgroups are indicated by color; the band represents the 95% confidence interval. (A-B) In both the Discovery and Replication Cohorts, MoCA scores decrease over time in the fast cognitive decline subgroup, while remaining stable in the slow cognitive decline subgroup. (C-D) In the Discovery Cohort, fast and slow cognitive decline subgroups do not differ in rate of motor change (UPDRS-III score) over time. However, in the Replication Cohort, the fast cognitive decline subgroup also experiences more rapid change in motor symptoms. (E) In the Replication Cohort, the fast cognitive decline subgroup has a faster rate of decline in the DRS score as well. (F) In the Replication Cohort, the fast cognitive decline subgroup has higher rates of incident MCI or dementia over 4 years of follow-up.

Rates of motor change (captured in the UPDRS-III scores over time) did not differ for Fast vs. Slow Cognitive Decline subgroups in the Discovery Cohort (**Figure 2C**). In the Replication Cohort, which had a longer disease duration at the time of plasma sampling, the Fast Cognitive Decline subgroup had a faster rate of motor decline (p<0.001, **Figure 2D**).

The UPenn-based Replication Cohort has a battery of neuropsychological assessments performed longitudinally, as well as longitudinal clinical cognitive consensus determination, as previously described.^35^ Since our method of assigning PD individuals to Fast vs. Slow Cognitive Decline subgroups was chosen for ease and applicability across all cohorts, it is possible that these groupings do not reflect meaningfully different trajectories. Thus, to verify our assignments of Fast vs. Slow Cognitive Decline subgroups, we compared rates of change in the Mattis Dementia Rating Scale-2 (DRS, a comprehensive measure of global cognition) for Fast vs. Slow Cognitive Decline subgroups in the UPenn cohort, using linear mixed effects models adjusted for age, sex, and disease duration. We found that the two subgroups differed significantly, with only the Fast Cognitive Decline subgroup showing decline in DRS scores over time (p<0.001, **Figure 2E**). We also compared rates of incident mild cognitive impairment (MCI) or dementia, as clinically determined, in the Fast vs. Slow Cognitive Decline subgroups in the UPenn PD cohort. In survival analyses, the Fast Cognitive Decline subgroup was more than twice as likely to develop incident MCI or dementia than the Slow Cognitive Decline subgroup (67.4% developed incident MCI or dementia in the Fast subgroup vs. 31.8% in the Slow subgroup, p = 0.006, **Figure 2F**).

Taken together, Fast Cognitive Decline subgroups assigned based on longitudinal MoCA scores comprised similar proportions of PD patients in both the Discovery and Replication cohorts, despite differences in clinical site and disease duration for these two cohorts. Moreover, subgroups assigned based on change in MoCA score had clinical significance; the Fast subgroup had greater rates of incident MCI or dementia. Thus, we used these Fast vs. Slow Cognitive Decline subgroup designations to develop and test biomarkers for cognitive decline in PD.

### Plasma proteins associating with rates of cognitive change in PD

In both the Discovery and Replication Cohorts, we measured levels of 940 plasma proteins using an aptamer-based platform.^30^ We tested each protein for association with Fast vs. Slow Cognitive Decline using a linear model adjusted for age, sex, and disease duration, generating an initial candidate list of nine proteins that associated with rates of cognitive decline at a p-value of <0.10, with the same directionality, in both the Discovery and Replication Cohorts (**Table 2 and Supplementary Table 2**).

**Table 2.** Top 9 SOMAScan Proteins. Nine proteins were associated with the rate of cognitive decline, in the same direction, at a p-value cut-off of 0.1 in both the Discovery and Replication Cohorts.

|  |  | Discovery Cohort |  | Replication Cohort |  |
| --- | --- | --- | --- | --- | --- |
| Protein Symbol | Protein Name | + Higher in Rapid Group<br>- Higher in Slow Group | P Value | + Higher in Rapid Group<br>- Higher in Slow Group | P Value |
| **MIA** | Melanoma Inhibitory Activity | + | 0.027 | + | 0.013 |
| **CRP** | C-Reactive Protein | - | 0.061 | - | 0.002 |
| **Albumin** | Albumin | + | 0.058 | + | 0.052 |
| **MIP.5** | Chemokine (C-C Motif) Ligand 15 | + | 0.032 | + | 0.037 |
| **FGF.5** | Fibroblast Growth Factor 5 | + | 0.048 | + | 0.051 |
| **AggreCAN** | AggreCAN | + | 0.049 | + | 0.037 |
| **ROBO3** | Roundabout Guidance Receptor 3 | + | 0.053 | + | 0.050 |
| **IL-4.sR** | Soluble IL-4 receptor | + | 0.015 | + | 0.022 |
| **RUXF** | Small Nuclear Ribonucleoprotein Polypeptide F | + | 0.083 | + | 0.036 |

These nine proteins, along with clinical variables of age, sex, and disease duration, were used to develop a logistic regression-based classifier to assign individuals to either the Fast or Slow Cognitive Decline subgroups in the Discovery Cohort. In five-fold cross-validation within the Discovery Cohort, the area under the receiver operating curve (AUC) for this model was 0.81. Applying the same logistic regression model to the Replication Cohort, which was never used to train the model, we obtained an AUC of 0.82, demonstrating that the model was not overfitted to the Discovery Cohort. In contrast, a model that used only the clinical variables of age, sex, and disease duration, and did not use any plasma biomarkers, only reached an AUC of 0.65 (**Figure 3A**). Finally, we considered including baseline MoCA score as an additional variable in the nine-protein model. However, this led to an increase in AUC for the Discovery Cohort (AUC = 0.86), with a decrease in AUC for the Replication Cohort (AUC = 0.77), suggesting overfitting with inclusion of this additional variable (**Supplementary Figure 1**).

**Fig 3.**
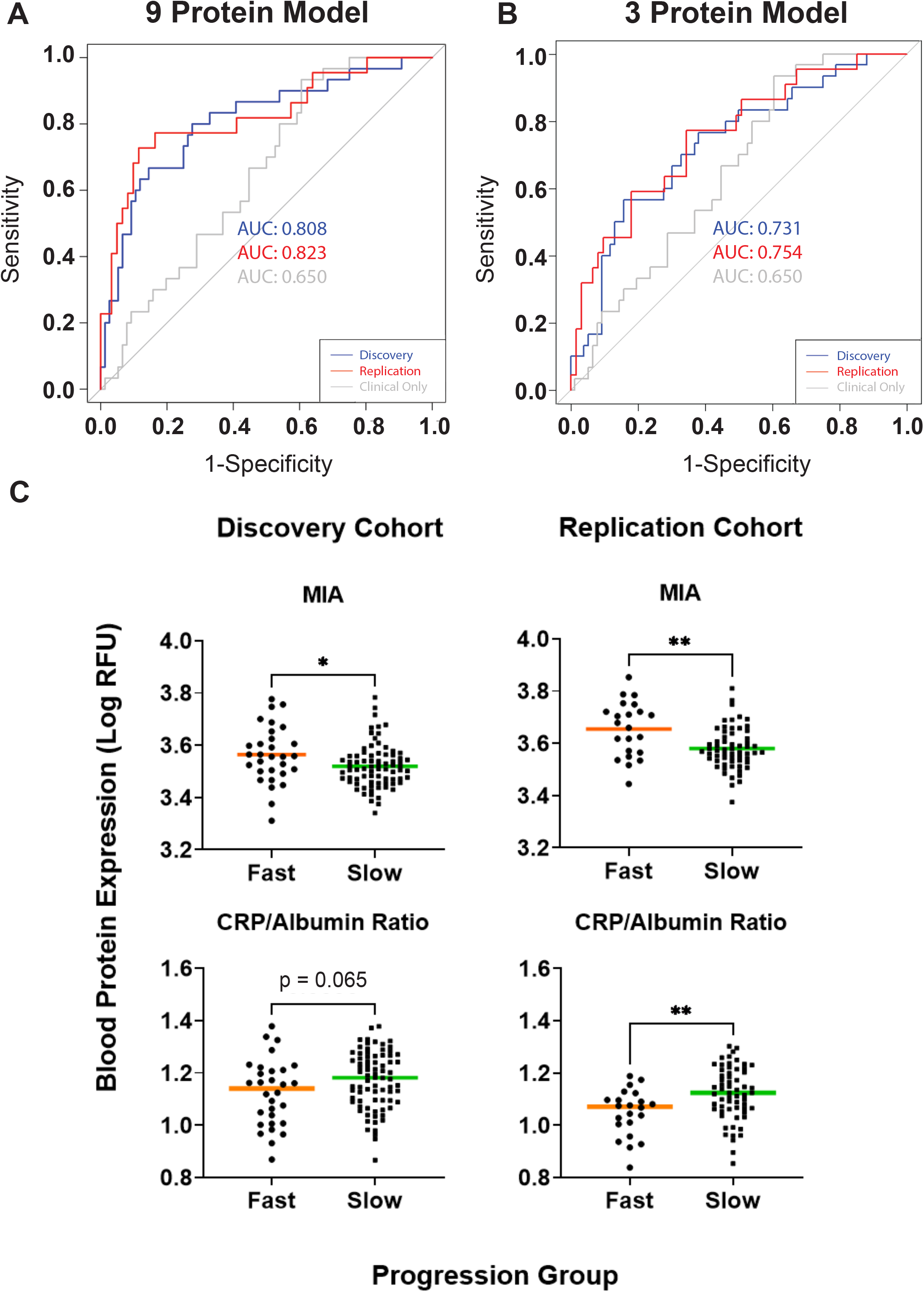
Identification of top biomarkers differentiating fast vs. slow cognitive decline subgroups in both Discovery and Replication Cohorts. (A-B) Performance characteristics of the logistic regression model for predicting whether an individual PD patient falls in the fast vs slow cognitive decline PD subgroup, trained using the measurements of all 9 proteins (panel A) or only 3 proteins (MIA, CRP, albumin, panel B), together with age, sex and disease duration. The model was trained using Discovery Cohort data and tested on the Replication Cohort. In the Discovery cohort, area under the receiver operating curve (AUC) was derived by five-fold cross-validation over 50 iterations. (C) Boxplots (median) showing the distribution of top biomarkers – MIA, CRP/Albumin Ratio levels – in log_10_ of RFU by PD cognitive decline subgroups. Mann-Whitney test was used to compare biomarker differences between fast vs. slow cognitive decline subgroups.

Among the nine proteins in the model were three proteins with high potential for downstream clinical biomarker translation based on existing translational uses and assay availability: C-reactive protein (CRP), albumin, and melanoma inhibitory activity protein (MIA). Specifically, CRP/albumin ratios are commonly used in the clinical setting to assess inflammatory status,^36-37^ and reagents for measuring MIA are readily available since MIA has been proposed as a biomarker for the common skin cancer melanoma.^38-39^ We thus evaluated the performance of a Fast vs. Slow Cognitive Decline classifier that required only age, sex, disease duration, and plasma levels of MIA, CRP, and albumin as input variables. As shown in **Figure 3B**, this three-protein model performed less well than the nine-protein model, but considerably better than the clinical variable-only model, with an AUC of 0.73 in the Discovery Cohort and 0.75 in the Replication Cohort. Moreover, in both the Discovery and Replication Cohorts, MIA plasma levels were higher in the Fast Cognitive Decline group (p = 0.026 for Discovery, p = 0.003 for Replication Cohort); the CRP/albumin ratio was lower in the Fast Cognitive Decline group in the Replication cohort (p = 0.004, with a similar trend in the Discovery Cohort (p = 0.065, **Figure 3C**).

### Validation of the top plasma biomarkers for cognitive decline

For downstream clinical translation, biomarkers need to be robust to changes in cohort and measurement platform. Thus, we sought to confirm our top biomarkers for cognitive decline in PD in an additional validation cohort, using a different method of measurement.

We first evaluated enzyme-linked immunosorbent assays (ELISAs) for quantitation of MIA and CRP in the plasma, using duplicate samples to compare measures obtained by ELISA vs. the original aptamer-based platform. MIA measures were moderately correlated (Pearson r = 0.79, **Figure 4A**) and CRP measures highly correlated (Pearson r = 0.98, **Figure 4B**) across platforms. In contrast, plasma albumin measures obtained on the aptamer-based platform did not correlate well with measures obtained with the widely-used bromocresol purple (BCP) assay (Pearson r = 0.29, **Figure 4C**).

**Fig 4.**
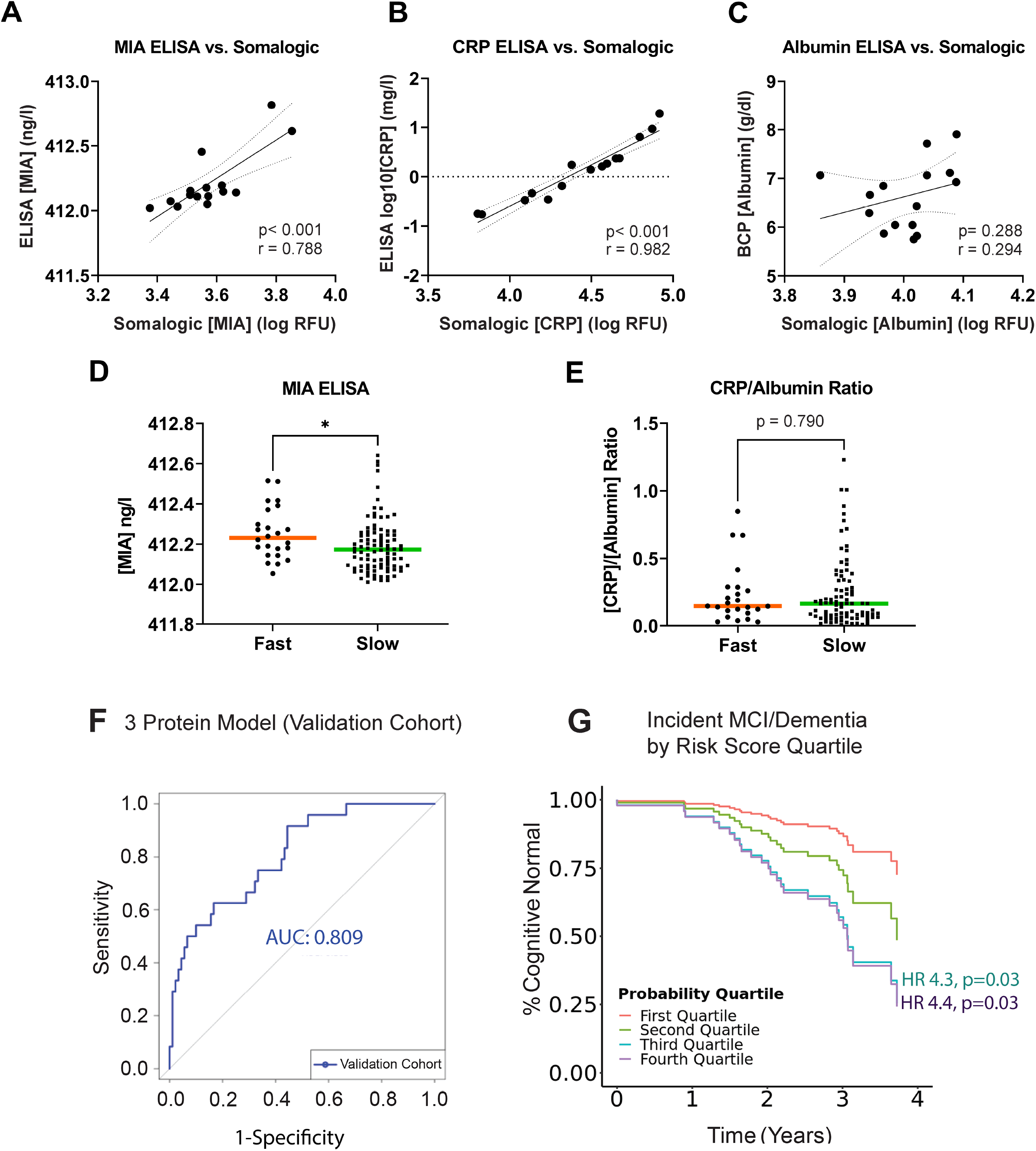
Validation of top biomarkers using alternative assays in the Validation Cohort. (A) Comparison of the values for 3 top biomarkers (MIA, CRP, Albumin) obtained on SOMAScan vs. Enzyme-linked immunosorbent (ELISA) or Bromocresol Purple (BCP) assay in 15 duplicate plasma samples. Pearson’s r is shown. (D-E) Boxplots (median) showing the distribution of MIA and CRP/Albumin ratio in the Validation Cohort by PD cognitive decline subgroups. Mann-Whitney test was used to compare the difference between subgroups. (F) Performance characteristics of the logistic-regression model (incorporating MIA, CRP, albumin, age, sex, and disease duration) for predicting fast vs slow cognitive decline subgroup in 118 PD patients from the Validation Cohort. (G) Time to incident MCI or dementia for PD patients in each quartile of risk score generated by the 6-parameter (3 protein, age, sex, disease duration) logistic regression model.

In a Validation Cohort of 118 longitudinally-followed PD patients from Penn (non-overlapping with the 83 Penn PD patients in the Replication Cohort), we measured plasma MIA and CRP by ELISA. Despite discrepancies between the aptamer-based platform and BCP measures for albumin, we also measured plasma albumin in the Validation Cohort with the BCP assay, as it is an assay commonly used in clinical settings, and our sample size of 15 for our cross-platform comparison might be underpowered.

Using the same criteria as the Discovery and Replication Cohorts, we first characterized PD individuals from the Validation Cohort as Fast vs. Slow Cognitive Decline, assigning 24/118 (20%) individuals to the Fast Cognitive Decline subgroup. In this group, plasma levels for MIA were significantly higher (p=0.022, **Figure 4D**), but the CRP/albumin ratio did not differ between Fast and Slow Cognitive Decline subgroups (**Figure 4E**).

Finally, we sought to validate a classifier for Fast vs. Slow Cognitive Decline in PD using these three plasma proteins – MIA, CRP, and albumin – as well as age, sex and disease duration in the 118-individual Validation Cohort. Despite differences in patient cohort and biomarker assays used, performance for this three-protein classifier was just as strong in the Validation Cohort, with an AUC of 0.81 (**Figure 4F**). Furthermore, when comparing a risk score based on this three-protein model to clinical outcomes, we found significantly greater rates of incident MCI or dementia among individuals with higher risk scores (**Figure 4G**), with a HR of 4.3 (p=0.03, 95% CI 1.15-15.9) for those in the third quartile of risk and a HR of 4.4 (p=0.03, 95% CI 1.19-16.5) for those in the highest quartile of risk score, compared to individuals in the lowest quartile of risk score.

Taken together, a three-protein predictor including MIA, CRP, and albumin, emerging from our unbiased screen of 940 plasma proteins, significantly enriched for PD individuals most likely to experience clinical cognitive decline in the near term. Moreover, results were not affected by differences in patient cohort or biomarker testing platform.

### MIA and cognitive decline in PD

Among the three proteins in our newly-validated risk predictor for cognitive decline in PD, CRP/albumin may indicate inflammatory status, but plasma MIA is harder to interpret biologically. We thus investigated this signal further in two ways. First, we sought to understand whether plasma MIA values, used alone, might risk-stratify PD patients in terms of future clinical cognitive decline. Second, we used Mendelian-randomization (MR)-based techniques to investigate MIA’s causal influence on the development of cognitive decline in PD.

Since both the UPenn-based Replication Cohort, and the UPenn-based Validation Cohort have clinical consensus diagnoses of normal cognition, MCI, or dementia, we asked whether individuals with higher plasma MIA were more likely to develop incident MCI or dementia over five years of follow-up. In both the Replication Cohort (**Figure 5A**), and the Validation Cohort (**Figure 5B**), we observed non-significant trends towards higher rates of incident MCI or dementia among PD patients with higher plasma MIA.

**Fig 5.**
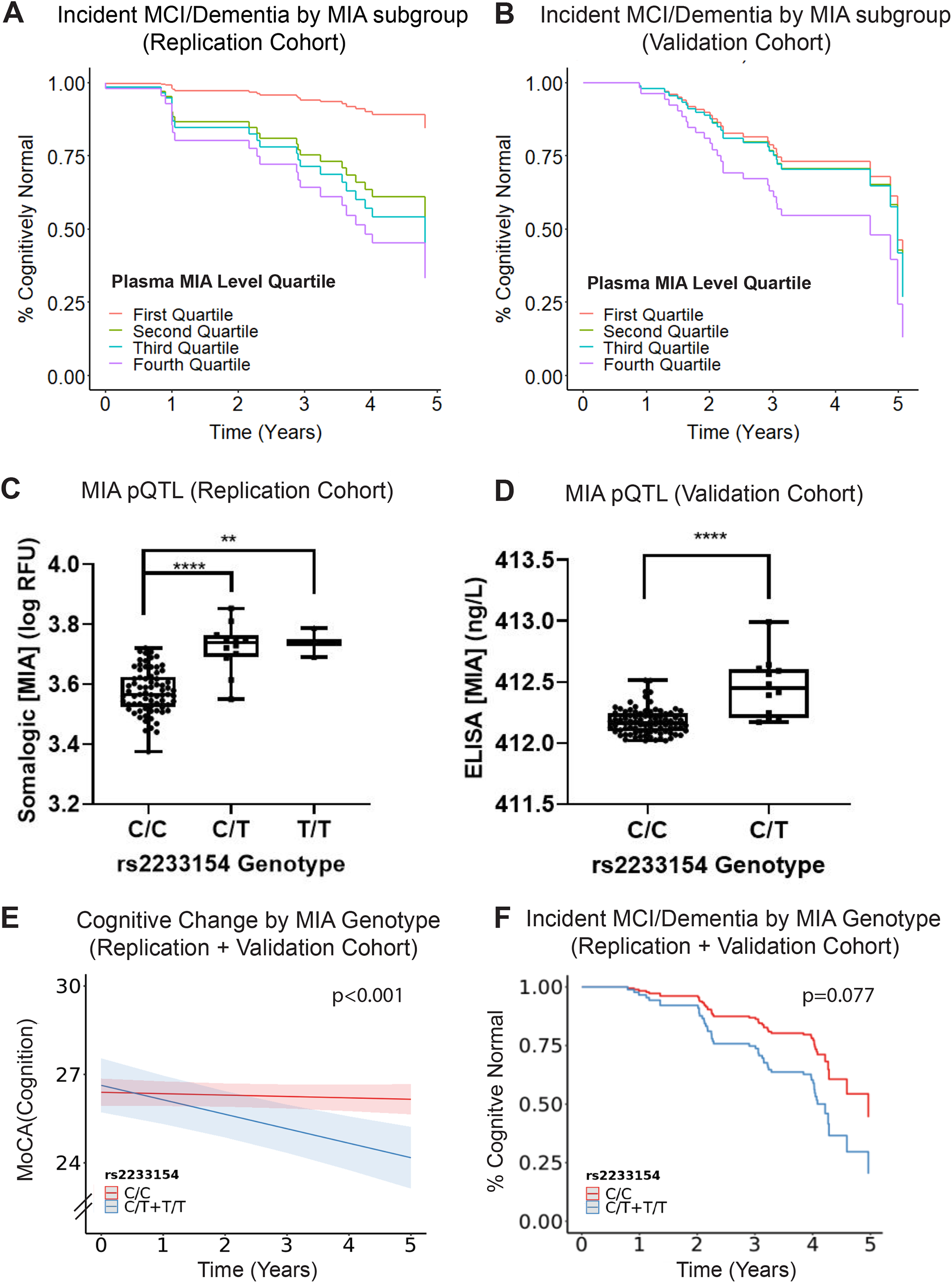
MIA as a novel blood biomarker for cognitive function decline in PD. (A-B) Cox proportional hazards model adjusted for age, sex and disease duration showing incident MCI or dementia rate for each quartile of baseline MIA measures in the (A) Discovery Cohort and (B) Validation Cohort over 5 years of follow-up. (C-D) Boxplot showing the association between genotypes at the *MIA* locus SNP rs2233154 and MIA expression in the plasma in the (C) Discovery Cohort and (D) Validation Cohort. There were no individuals with the TT genotype in the Validation Cohort. (E) Effect of rs2233154 genotype on longitudinal MoCA performance assessed using linear-mixed effects models adjusting for age, sex and disease duration. (F) Cox proportional hazards model adjusting for age, sex and disease duration shows a differential rate of incident MCI or dementia comparing carriers of different rs2233154 genotypes over 5 years of follow-up.

For our MR-based analyses of causal inference for MIA’s role in PD cognitive decline, we first identified expression quantitative trait locus (eQTL) single nucleotide polymorphisms (SNP) for *MIA* mRNA expression from the Genotype-Tissue Expression database (GTEx).^34^ Carriers of one or more minor (T) alleles at rs2233154 demonstrated higher *MIA* expression in multiple tissues, including several brain regions (cortex, amygdala, hypothalamus, pituitary), the tibial nerve, several vascular tissues (aorta, left ventricle of the heart), and several gastro-intestinal tissues (sigmoid colon, esophagus) in GTEx data. Thus, we compared levels of plasma MIA among carriers of different rs2233154 genotypes. As shown in **Figures 5C and 5D**, rs2233154 genotypes associated significantly with MIA plasma protein levels in both the Replication Cohort (rs2233154^CC^ ^vs. CT^ p = < 0.0001, rs2233154^CC^ ^vs. TT^ p = 0.0076) and the Validation Cohort (rs2233154^CC^ ^vs. CT^ p = < 0.0001).

We next asked whether carriers of rs2233154 genotypes differed in rates of cognitive decline. Since the Replication and Validation Cohort are both UPenn-based, and they were both assessed for cognition and assigned cognitive diagnoses in the same way, we combined them for a total set of n=180 PD individuals for these analyses.

In a linear mixed-effects model adjusted for age, sex, disease duration, and baseline MoCA score, rs2233154 genotypes associated significantly with rates of decline in the MoCA (p = 0.0004, **Figure 5E**), with carriers of one or more T alleles declining more rapidly. Specifically, for each additional T allele carried, MoCA scores declined by an additional 0.445 points/year, compared to trajectories for individuals without T alleles. Moreover, in a Cox proportional hazards model adjusting for age, sex, and disease duration, rs2233154 genotypes trended towards association with incident clinical MCI or dementia, with the same direction of effect (HR 1.8 for carriers of one or more T alleles [95% CI 0.94-3.4, p = 0.077], **Figure 5F**).

Taken together, these analyses suggest that MIA may not only serve as a biomarker for, but also play a causal role in, the development of cognitive decline in PD.

## DISCUSSION

In this study, we investigated three different PD cohorts from two different clinical sites, comprising a total of 309 longitudinally-followed individuals with PD, in order to discover, replicate, and validate biomarkers predictive of cognitive decline. In 191 individuals with PD, we first screened 940 plasma proteins for association with the rate of change in MoCA scores, nominating top proteins for downstream investigation. We then developed two models – each featuring multiple proteins as well as clinical variables – and tested them for ability to predict the rate of cognitive decline. For the simpler model, requiring measures of just three proteins, we developed alternate assays for these proteins and tested their performance in predicting both decline in cognitive test scores and incident MCI or dementia in an additional 118 PD individuals, demonstrating that our model can substantially enrich for individuals at high risk for rapid cognitive decline. Finally, for one of the novel proteins discovered in this process, MIA, we present evidence for a causal role from Mendelian randomization analyses.

Our findings have relevance for clinical prognostication as well as planning of clinical trials aimed at modifying the rate of cognitive decline in PD. Specifically, while prior studies have addressed the question of cognitive decline in PD, most studies that incorporate biochemical biomarkers focus on demonstrating significant associations between a given biomarker and cognitive trajectory,^2,4,22^ rather than developing tools that might be applied at an individual level for risk stratification. A few studies have built predictors applicable to individuals. For example, Liu *et al*. reported a clinicogenetic predictor with an AUC of 0.85 for prediction of incident cognitive impairment within 10 years.^40^ More recently, Tang *et al*. used both clinical and radiographic data from the Parkinson’s Progression Markers Initiative (PPMI) cohort to predict time to cognitive progression, employing a training-test design similar to the one used in our study and finding that clinical variables, in particular, were highly predictive of incident MCI.^41^ Neither the Tang or Liu reports, however, focus on biochemical biomarkers. As such, our findings contribute to knowledge in the field with respect to what plasma proteins may add to ability to predict individual cognitive decline. Given the routine use of biochemical biomarkers for risk stratification in other areas of medicine (*e*.*g*. lipid levels in cardiology, tumor markers in oncology), development and validation of easily-accessed protein biomarkers that can be used at an individual level in PD is a priority.^27^

Our study also yields insight into the mechanisms that may lead to cognitive decline in PD. First, our finding that plasma CRP levels may be predictive of rate of cognitive change in PD should be considered in light of prior reports demonstrating that CRP levels are higher in PD compared to neurologically normal controls.^42^ In both our Discovery and Replication cohorts, however, PD individuals with higher CRP levels experience *less* subsequent cognitive decline. Thus, our CRP findings contextualize the prior literature in several ways. They show that CRP emerges from a screen of nearly 1000 proteins as an informative biomarker of disease trajectory, adding confidence to a signal that has largely been evaluated in candidate-protein studies. Additionally, they suggest that a more nuanced view is needed with respect to whether therapeutic approaches aimed at reducing inflammation – and consequently CRP levels – would be beneficial in PD, since higher-CRP individuals tended to maintain their cognitive status in our study. Second, our study highlights plasma MIA as a biomarker predicting cognitive decline in PD, with evidence to support a causal role for MIA-related pathways in the development of these cognitive features. MIA is a secreted protein most well-characterized for its role as a biomarker in the skin cancer melanoma. Known to be highly expressed in malignant melanocytes, MIA has been used as a blood-based biomarker in cancer, with higher levels indicative of risk for metastatic melanoma.^43,44^ To our knowledge, MIA has not been previously linked to PD. Intriguingly, however, melanoma and PD have long been known to co-occur at rates that are significantly higher than expected,^45^ and melanoma patients without PD have been reported to have a 10.5-fold relative risk of death from metastatic melanoma compared to melanoma patients with PD.^46^ Taken together, both these epidemiological studies and our current causal inference analysis of MIA support mechanistic investigation of MIA-related pathways in PD.

Strengths of this study include (1) the large-scale screening of 940 plasma proteins as potential biomarkers predicting cognitive decline in PD, through which novel leads might be discovered, (2) the discovery-replication-validation design, ensuring that no single cohort, method of ascertaining cognitive impairment, or protein measurement platform may be responsible for our findings, and (3) the incorporation of analyses aimed at understanding whether MIA as a top biomarker might be *causal* for the development of cognitive decline in PD. Few reported biomarker studies in PD have incorporated protein screening at the scale used here, with even fewer incorporating detailed longitudinal follow-up of patients for clinically-relevant outcomes. Moreover, while some discovery screening studies offer promising leads,^47^ to date, these biomarker candidates largely lack replication.

Limitations of our study should also be considered. First, while our study incorporated three different cohorts of PD patients, with each phase designed to replicate (or fail to replicate) the findings of the prior phase, two-thirds of the PD individuals studied here were from one site (UPenn). Future studies evaluating our cognitive decline predictor in larger, multi-site cohorts at various stages of disease are needed to translate our findings into the most useful real-world applications. Second, some findings were only incompletely replicated across phases of our study. In particular, the CRP/albumin ratio, lower in the Slow Cognitive Decline subgroup in the UPenn-based Replication Cohort, did not differ comparing Slow vs. Fast Cognitive Decline subgroups in the UPenn-based Validation Cohort. This inconsistency might be due to biological noise. Alternately, the lack of replication might stem from the measurement methods used in the Validation Cohort, especially as plasma albumin measures did not correlate well for the SOMAScan vs. BCP assay. Nonetheless, it is reassuring that performance for the three-protein predictor remained as high in the Validation Cohort (AUC 0.80) as in the other phases of our study. Third, while both our nine-protein model and our three-protein model show moderate performance (AUC 0.73-0.82), and certainly improve upon the ability to make predictions based on clinical variables alone (AUC 0.65), ability to separate Fast vs. Slow Cognitive Decline groups is not perfect. It is likely that the arbitrary division of PD individuals into two cognitive decline groups based on change in MoCA scores precludes our ability to see the fuller separation one might expect for groups that are truly biologically distinct. That said, our goal in this study was to create a robust, easy-to-use tool that might risk-stratify PD patients at the individual level. We point to the fact that PD individuals in the highest 25% of risk score were 4.4 times more likely to develop incident MCI or dementia in four years than those in the lowest 25% of risk score.

In summary, we present our findings from a study of 309 longitudinally-followed PD individuals, where, starting from 940 plasma protein candidates, we develop a risk predictor for cognitive decline in a four-year window. We find that a risk score based only on age, sex, and plasma values of MIA, CRP, and albumin identifies a subgroup of PD individuals 4.4 times more likely to develop cognitive decline in the near term, regardless of cohort studied, cognitive measure used, or method for biomarker measurement. We furthermore link MIA causally to development of cognitive impairment through Mendelian randomization. Taken together, our study offers an easy-to-use tool for risk stratification for future cognitive impairment in PD individuals, as well as a new lead for mechanistic investigation. The development of molecular tools – such as the risk score calculator presented here – enables “precision medicine” approaches to the care of PD patients. Moreover, the emergence of MIA from a 940-protein screen illustrates an approach for deriving targets for downstream mechanistic experiments based on biochemical profiling of patient-derived biofluids.

## Data Availability

All data produced in the present study are available upon reasonable request to the authors

## ACKNOWLEDGMENTS

We thank Maria Diaz-Ortiz for technical assistance. We additionally thank our patients and their families for their generosity in contributing to this research. We thank the NINDS PDBP and the BioSEND biorepository for assistance with samples.

This research was supported by the NIH (RO1 NS115139, U19 AG062418, P50 NS053488), and a Biomarkers Across Neurodegenerative Diseases (BAND) grant from the Michael J. Fox Foundation/Alzheimer’s Association/Weston Institute. Alice Chen-Plotkin is additionally supported by the Parker Family Chair, the AHA/Allen Institute Brain Health Initiative, and the Chan Zuckerberg Initiative Neurodegeneration Challenge.

## AUTHOR CONTRIBUTIONS

Author contributions to this work are as follows: ACP designed the study, with substantial input from JS. NA, RZ, RTS, TLU, MP, TFT, VVD, RBD, and DW acquired the data. JS, NA, SXX, MP and ACP analyzed the data. JS, NA, and ACP drafted the majority of the manuscript, and all authors edited and approved the final manuscript.

## SUPPLEMENTARY MATERIALS

**Shen et al, “Unbiased screen of 940 proteins identifies MIA, CRP, and albumin as plasma biomarker predicting cognitive decline in Parkinson’s Disease.”**

## Supplementary Methods

### Cohorts

#### University of Texas Southwestern (UTSW) Discovery Cohort

The NIH-NINDS PDBP is the parent study into which the UTSW Discovery Cohort was enrolled. All individuals were followed on an annual basis; cognitive testing by the Montreal Cognitive Assessment (MoCA) was obtained longitudinally, and blood was collected under standard operating procedures.^1^ Out of 115 PD individuals for which SOMAScan data was previously obtained, 108 individuals had a MoCA score of 19 or greater at the time of plasma sampling, as well as longitudinal cognitive testing data, and these individuals were included in the UTSW Discovery Cohort.^1,2^ The maximum follow-up time for the UTSW Discovery Cohort is four years (interquartile range [IQR] is 1.0-3.0 years with a median follow-up of 3.0 years). The UTSW institutional review board (IRB) approved study protocols, and all participants were consented for the study.

#### UPenn Replication Cohort and UPenn Validation Cohort

The parent study of both the UPenn Replication Cohort and the UPenn Validation Cohort is the Clinical Core of the UPenn NIA U19 (Center on alpha-synuclein strains in Alzheimer’s disease and related dementias at the Perelman School of Medicine at the University of Pennsylvania, formerly the Morris K. Udall Center). Over the past 13 years, the Clinical Core of the UPenn NIA U19 has recruited PD subjects to participate in a longitudinal study that includes (1) serial assessment of cognition, and (2) biofluid sampling for biomarker and DNA analysis, with approximately 180 active participants at any given time. 201 PD participants from the UPenn NIA U19 cohort had plasma samples available for biomarker testing, were non-demented at the time of plasma sampling, and had at least 4 years of cognitive testing and determination of cognitive diagnosis by clinical consensus after the time of sampling. Of these 201, SOMAscan measures were previously obtained on 83, and these individuals formed the UPenn Replication Cohort. The remaining 118 PD individuals comprised the UPenn Validation Cohort. Written informed consent was obtained at study enrollment, and the UPenn IRB approved study protocols. Although the UPenn NIA U19 PD participants have up to 13 years of follow-up, for our protein biomarker analyses, we used cognitive data from only four or five years of follow-up after plasma sampling (time period is indicated in the text) in this study for two reasons. First, we sought to match the Replication Cohort duration of follow-up to that of the Discovery Cohort. Second, as the vast majority of PD individuals will develop dementia over their disease course, we reasoned that the development of incident MCI or dementia in a fixed, relatively short period of time is more clinically meaningful for the purposes of our survival analyses.

### Statistical Analyses

#### Nomination of proteins that differentiated PD individuals with Fast vs. Slow Cognitive Decline

A linear regression model was used to identify proteins whose plasma concentration associated significantly with cognitive subgroup (fast vs. slow cognitive decline) in both the Discovery and Replication Cohorts. Age and disease duration at the time of plasma sampling, sex, as well as cognitive subgroup (fast vs. slow) were included as independent variables, while the individual protein concentrations (n=940) were used as outcome variables. Proteins that associated with cognitive decline subgroup in both the Discovery and Replication Cohorts (1) at a nominal p-value of 0.1 or less and (2) with the same direction of association in both cohorts were selected for downstream investigation.

#### Development of models to predict fast vs. slow cognitive decline

The nine proteins selected for their nominal association with fast vs. slow cognitive decline subgroups were incorporated in a logistic regression model for binary classification. Specifically, our model predicted whether an individual would belong to the fast or slow cognitive decline subgroup based on measures for these nine proteins, age, sex, and disease duration, using the Discovery Cohort to train the model (obtaining weights for all variables). We then tested the exact model developed in the Discovery Cohort on the Replication Cohort, in order to evaluate its performance in a cohort whose data were never used to train the model. Performance was assessed with Receiver Operating Curve (ROC) analyses; we obtained area under the curve (AUC) measures by fivefold cross-validation over 50 iterations. In addition, we also developed a simpler model incorporating only three top proteins, age, sex, and disease duration, using the same methods as for the nine-protein model.

#### Linear mixed-effect model analyses

Linear mixed-effects models were used to evaluate the effect of biomarker levels on change in cognitive scores over time. A random intercept was introduced to the mixed effects model to account for correlations among multiple repeated measurements. Fixed effects included the interaction between plasma protein measure stratified by quartile and time, baseline MoCA score, age at sample collection, sex, and disease duration.

#### Cox proportional hazard models and survival analyses

To understand whether biomarkers associated with clinical outcomes, we performed survival analyses based on whether individuals developed incident MCI or dementia (converting from normal cognition to MCI, normal to dementia, or MCI to dementia, as determined by expert clinical consensus). In some analyses, individuals were binned into quartiles based on (1) measures for candidate protein biomarkers or calculated risk score for fast cognitive decline. Hazard ratios for development of incident MCI or dementia were then calculated for subgroups of patients as indicated in the text. For comparison of subgroups based on quartiles of protein measures, we performed Cox proportional hazards analyses, adjusting for age at sample, sex and disease duration. For comparison of subgroups based on calculated risk score for fast cognitive decline, we did not adjust for covariates, as clinical variables were already incorporated in the model for calculating cognitive decline risk scores.

---

**Supplementary Figure 1.**
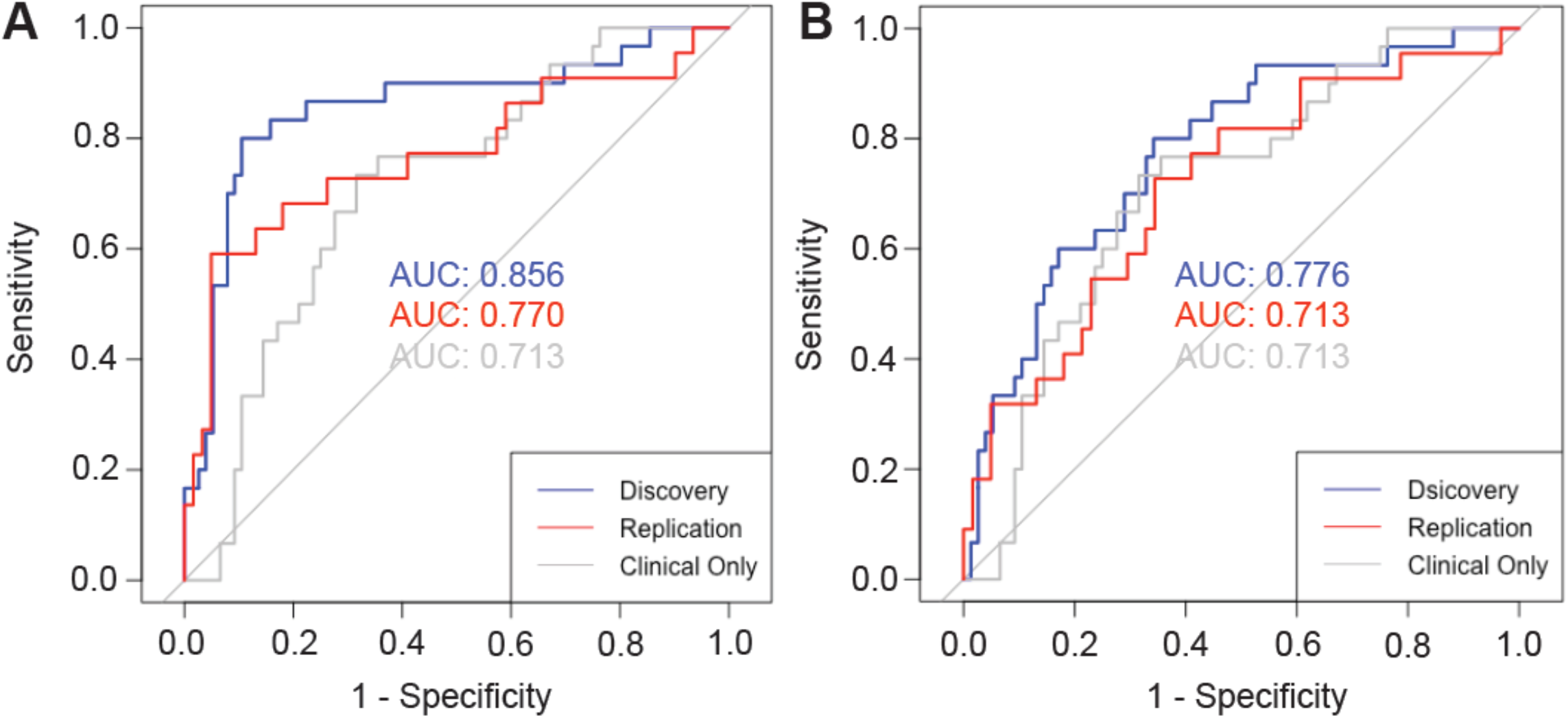
(A-B) Performance characteristics of the logistic regression model for predicting whether PD patient falls in the fast vs. slow cognitive decline subgroup, trained using the measurements of 9 proteins (panel A) or only 3 proteins (MIA, CRP, Albumin, panel B) together with age, sex, disease duration, and baseline MoCA.

